# Safety of Bio-Interventional Uveoscleral Outflow Enhancement: Twelve-Month Post-Operative Results from 395 Eyes in the CREST Study

**DOI:** 10.64898/2026.11.12.26367928

**Authors:** Douglas Rhee, James Lewis, Ticiana De Francesco, Kiper Nelson, Tsontcho Ianchulev, Steven R Sarkisian

**Affiliations:** Case Western Reserve University School of Medicine, Cleveland, Ohio; SightMd, Elkins Park, PA, USA; University of Utah John A. Moran Eye Center, Salt Lake City, UT; Southern Eye Center, Hattiesburg, MS, USA; New York Eye and Ear Infirmary of Mount Sinai, New York, NY; Oklahoma Eye Surgeons, Oklahoma City, OK, USA

**Keywords:** allograft, bio-reinforced cyclodialysis, glaucoma, safety, scleral allograft, uveoscleral outflow

## Abstract

**Purpose:** To characterize 12-month ocular safety of bio-interventional uveoscleral outflow enhancement in open-angle glaucoma (OAG).

**Design:** Prospective, multicenter real-world evidence study (CREST; NCT05506423).

**Participants:** Adults with OAG treated by bio-interventional uveoscleral outflow enhancement with acellular scleral allograft spacers (AlloFlo™, Iantrek, Inc.), standalone or with phacoemulsification, reaching 12 months of follow-up.

**Methods:** Adverse events were captured prospectively at each protocol-specified visit against pre-defined definitions; data were analyzed at the eye level.

**Main Outcome Measures:** Pre-specified ocular safety endpoints: adverse events, clinically significant hypotony, sustained intraocular pressure (IOP) elevation ≥ 10 mmHg beyond 30 days, allograft migration, corneal endothelial decompensation, secondary incisional glaucoma surgery, and endothelial cell density.

**Results:** Of 395 eyes (mean age 72.8 years; 52% female), 267 were combined with phacoemulsification (Cohort 1) and 128 standalone (Cohort 2). Medicated IOP decreased from 19.1 to 14.5 mmHg (p < 0.001) and medications from 1.9 to 1.2, with 42% medication-free. Sustained IOP elevation occurred in 34 eyes (8.6%), BCVA loss ≥ 2 lines in 15 (3.8%), hyphema > 2 mm in 6 (1.5%) and hypotony in 1 (0.3%). Two allograft spacers entered the anterior chamber in one eye and were successfully repositioned intraoperatively. Serious events occurred in 12 eyes (3.0%). In the N=91 eyes with paired specular microscopy data, endothelial cell density changed –6.5% (–8.7%, –4.4%). There were no reports of endothelial decompensation, endophthalmitis, or sympathetic ophthalmia.

**Conclusions:** Through 12 months no endothelial decompensation, endophthalmitis or sympathetic ophthalmia occurred, and most events were self-limiting or medically managed. Endothelial cell loss tracked concurrent cataract surgery rather than the allograft, though that subgroup is small. Follow-up remains shorter than the interval over which an earlier supraciliary microstent diverged from controls.

**Trial Registration:** ClinicalTrials.gov (NCT05506423.)

## Introduction

Open-angle glaucoma (OAG) is a leading cause of irreversible blindness worldwide, affecting more than 80 million people globally [1]. While IOP reduction remains the cornerstone of glaucoma management, a substantial proportion of patients require surgical intervention when medical therapy fails to achieve adequate IOP control. The surgical management of OAG has evolved considerably with the introduction of minimally invasive glaucoma surgery (MIGS), yet a fundamental limitation constrains the efficacy of trabecular-based procedures: the IOP-lowering effect of canal-based MIGS is limited by resistance in distal outflow pathways and the episcleral venous pressure, creating a physiologic IOP floor of approximately 16 mmHg [2].

Targeting the uveoscleral outflow pathway offers a physiologically distinct route for aqueous drainage and may overcome this limitation. The uveoscleral pathway accounts for 35%–50% of total aqueous outflow in healthy eyes and is largely independent of episcleral venous pressure [3]. Surgical access through cyclodialysis has been attempted since Heine’s original description in 1905, but the technique fell out of favor due to unpredictable cleft closure, rebound hypertension, and hypotony [4]. More recently, synthetic supraciliary microstents have demonstrated the hydraulic efficacy of the pathway with significant and sustained IOP reduction, though long-term safety has been challenged by reports of corneal endothelial cell loss associated with anterior-chamber hardware protrusion [5, 6].

Bio-reinforced cyclodialysis with AlloFlo™ (Iantrek, Inc.) uses a biocompatible acellular allogeneic scleral allograft scaffold to reinforce the cyclodialysis cleft from within, without supraciliary or intracameral hardware protrusion. The scaffold is biologically homologous to the surrounding sclera and is intended to support cellular repopulation, rather than the chronic foreign-body response associated with synthetic supraciliary devices.

Despite the expansion of MIGS evidence, the quality of adverse event reporting across MIGS studies has been inconsistent. A systematic review of 21 MIGS randomized controlled trials found that, on average, only 10 of 16 CONSORT for Harms checklist items were fulfilled, and only 4 of 21 trials provided pre-specified definitions of adverse events [7]. Pre-specified, protocol-defined adverse event criteria are therefore methodologically important for characterizing the safety of novel procedures.

Early clinical experience with bio-interventional uveoscleral outflow enhancement has shown encouraging efficacy and safety profile [8–10]. Given the relative novelty of the procedure, systematic characterization of its ocular safety in a large, prospective, multicenter cohort is warranted. This study reports the surgical safety of bio-interventional uveoscleral outflow enhancement across 395 eyes enrolled in the CREST Study with at least 12 months of follow-up. To date, this is the largest prospective safety assessment performed for this procedure.

## Methods

### Study Design

The CREST Study (NCT05506423) is a multicenter, observational, real-world evidence study, in which patients with OAG who underwent bio-interventional uveoscleral enhancement surgery with AlloFlo Uveo are prospectively followed through 24 months postoperatively. CREST is conducted in accordance with the Declaration of Helsinki. The study has received approval from the relevant Institutional Review Boards (IRBs) for all participating investigators. All participants provided written informed consent prior to enrollment.

The present analysis includes all eyes that completed at least 12 months of postoperative follow-up at the time of the data cutoff. Two cohorts were defined per-protocol, based on whether the procedure was performed in conjunction with cataract surgery (Cohort 1) or as a standalone procedure (Cohort 2). No additional inclusion or exclusion criteria beyond the study’s four eligibility requirements (confirmed diagnosis of OAG, procedure attempted or completed, provided signed written consent, and ability and willingness to follow study instructions) were applied. The CREST Protocol contains no exclusion criteria.

### Surgical Procedure

One, two, or three units of processed, acellular scleral allograft (AlloFlo™, Iantrek, Inc.) were deployed into the supraciliary space to maintain cyclodialysis cleft patency and serve as a porous conduit for aqueous humor drainage into the uveoscleral pathway. The bio-reinforced cyclodialysis procedure was performed using an ab-interno approach under gonioscopic visualization. Elongated 5 mm × 500 µm acellular scleral allograft bio-scaffolding constructs are prepared and introduced into the AC (Figure 1) under gonio-visualization (Figure 2). The reinforcing spacers are deployed at the endoscleral surface and adjacent to the edges of the cyclodialysis. The treating surgeon has discretion to use the number of allograft spacers needed to achieve the desired sustained cleft opening.

Components do not protrude the drainage angle or anterior to Schwalbe’s line, and placement is confirmed under direct visualization with the bio-tissue spacers deployed flush with the iris root. Any residual viscoelastic is evacuated with irrigation or irrigation/aspiration. Patients were prescribed a surgeon’s choice of postoperative regimen of topical antibiotics and corticosteroids with taper.

### Safety Endpoints

The primary focus of this analysis is ocular adverse events through 12 months of follow-up. Intraoperative ocular adverse events captured on the operative-day case report form included allograft malposition or dislodgement at the time of surgery, hyphema requiring intraoperative management, and unplanned conversion or termination of the procedure.

Anterior chamber cells and flare were graded on a 0 to 3+ scale at each protocol visit and, as for corneal edema, are reported at visits occurring ≥ 30 days after surgery, excluding the transient inflammation expected in the immediate postoperative period.

Each event was graded for severity (mild, moderate, severe) and for seriousness. An event was recorded as serious if it resulted in death, was life-threatening, required inpatient hospitalization or prolongation of existing hospitalization, resulted in persistent or significant disability or incapacity, or required intervention to prevent permanent impairment or damage. Relatedness was recorded on four separate fields, according to the investigator’s assessment: relationship to any of the allograft, delivery system, intraocular lens, or other materials. Both severity and relatedness were assigned by the treating site investigator at the time of the event. Investigators were not masked, and no independent or blinded adjudication committee reviewed causality. Relatedness is therefore reported throughout as an investigator-assessed, rather than adjudicated determinations.

### Statistical Analysis

Safety endpoints are reported as proportions with 95% confidence intervals (95% CIs) computed using the Wilson score method. Longitudinal IOP change was analyzed using the Wilcoxon signed-rank test for paired baseline and 12-month data, as IOP differences did not follow a normal distribution (Shapiro-Wilk p < 0.001). Medication burden change was similarly analyzed using the Wilcoxon signed-rank test. Change in corneal endothelial cell density was analyzed using a two-sided paired t-test of the within-eye percent change from baseline, and paired binary outcomes such as the proportion of eyes on no IOP-lowering medication were compared using McNemar’s test. The two cohorts were compared on endothelial cell change using a two-sample t-test; no adjustment was made for multiple comparisons. Efficacy results are presented as means with 95% CI or ± standard deviation (SD) where appropriate. No statistical comparison was made between this cohort and any external dataset. Published rates referenced in the Discussion sections are provided for context only. Analyses were performed using R version 4.5.3 (R Foundation for Statistical Computing).

## Results

### Demographics and Baseline Characteristics

A total of 395 eyes of 298 patients completed at least 12 months of follow-up and were included for analysis. Mean age was 72.8 (8.6) years; 207 eyes (52%) were from female patients, and 272 eyes (69%) were from white patients (Table 1). The cohort reflected a real-world OAG population with moderate-to-advanced disease: mean baseline medicated IOP was 19.1 (18.5, 19.7) mmHg, patients were prescribed a mean of 1.9 (1.5) IOP-lowering medications at baseline (33% on ≥ 3), and 24% of eyes were designated by the treating physician as candidates for bleb-forming surgery prior to the study procedure.

**Table 1.** Summary of Demographics and Baseline Disease Characteristics (N=395)

| **Characteristic** | **Cohort 1<br>(N=267)** | **Cohort 2<br>(N=128)** | **Total<br>(N=395)** |
|---|---|---|---|
| **Demographics** |  |  |  |
| Age, mean (±SD) years | 72.9 (7.5) | 72.8 (10.5) | 72.8 (8.6) |
| Female, n (%) | 143 (54%) | 64 (50%) | 207 (52%) |
| White, n (%) | 193 (72%) | 79 (62%) | 272 (69%) |
| **Baseline Disease Characteristics** |  |  |  |
| IOP, mean (95% CI) mmHg | 18.1 (17.5, 18.7) | 21.3 (19.9, 22.6) | 19.1 (18.5, 19.7) |
| IOP-lowering medications, mean (±SD) | 1.4 (1.3) | 3.0 (1.5) | 1.9 (1.5) |
| 0 medications, n (%) | 64 (24%) | 9 (7%) | 73 (18%) |
| 1 medication, n (%) | 110 (41%) | 14 (11%) | 124 (31%) |
| 2 medications, n (%) | 44 (17%) | 25 (20%) | 69 (17%) |
| ≥ 3 medications, n (%) | 49 (18%) | 80 (63%) | 129 (33%) |
| Visual field MD, mean (±SD) dB | −7.0 (7.1) | −10.6 (8.0) | −8.4 (7.6) |
| Pseudophakic, n (%) | 0 (0%) | 120 (94%) | 120 (30%) |
| Bleb-forming candidate, n (%)<sup>a</sup> | 0 (0%) | 96 (75%) | 96 (24%) |
| **Prior Glaucoma Procedures** |  |  |  |
| Mean number of prior procedures (±SD) | 0.2 (0.6) | 1.8 (1.6) | 0.7 (1.3) |
| 0 prior procedures, n (%) | 229 (86%) | 30 (23%) | 259 (66%) |
| 1 prior procedure, n (%) | 30 (11%) | 29 (23%) | 59 (15%) |
| 2 prior procedures, n (%) | 3 (1%) | 30 (23%) | 33 (8%) |
| ≥ 3 prior procedures, n (%) | 5 (2%) | 39 (30%) | 44 (11%) |
| Prior trabecular MIGS or laser, n (%) | 36 (13%) | 88 (69%) | 124 (31%) |
| Prior bleb-forming surgery, n (%) | 2 (1%) | 33 (26%) | 35 (9%) |

CI, confidence interval; IOP, intraocular pressure; MD, mean deviation; MIGS, minimally invasive glaucoma surgery; SD, standard deviation.
a. Designated by treating physician as candidate for bleb-forming surgery at time of study procedure if bio-reinforced cyclodialysis were not available.

Most eyes (267 [68%]) underwent combined cataract surgery and bio-reinforced cyclodialysis (Cohort 1), and 128 (32%) underwent standalone bio-reinforced cyclodialysis (Cohort 2). Most eyes (211 [54%]) received two allograft spacers; 162 (41%) received a single spacer, and 21 (5%) received three or more. Allograft count was not recorded in one eye.

### Ocular Adverse Events

Pre-specified adverse events are summarized in Table 2. The most common were sustained IOP elevation ≥ 10 mmHg above baseline occurring > 30 days postoperatively (34 eyes, 8.6%) and BCVA loss of ≥ 2 lines at ≥ 90 days postoperatively (15 eyes, 3.8%). IOP elevations were managed with topical or oral therapy, office laser, or secondary incisional surgery.

**Table 2.**
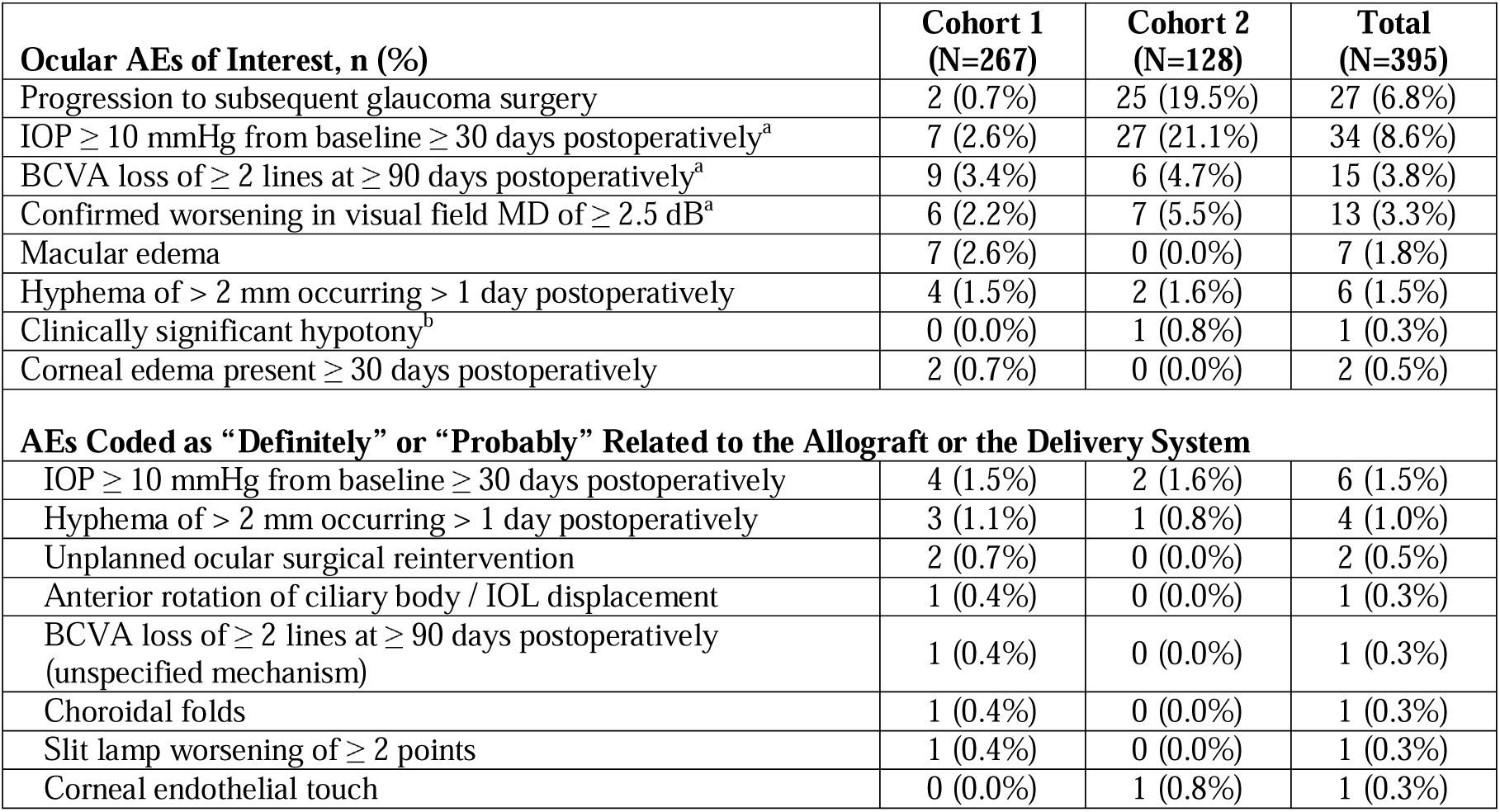
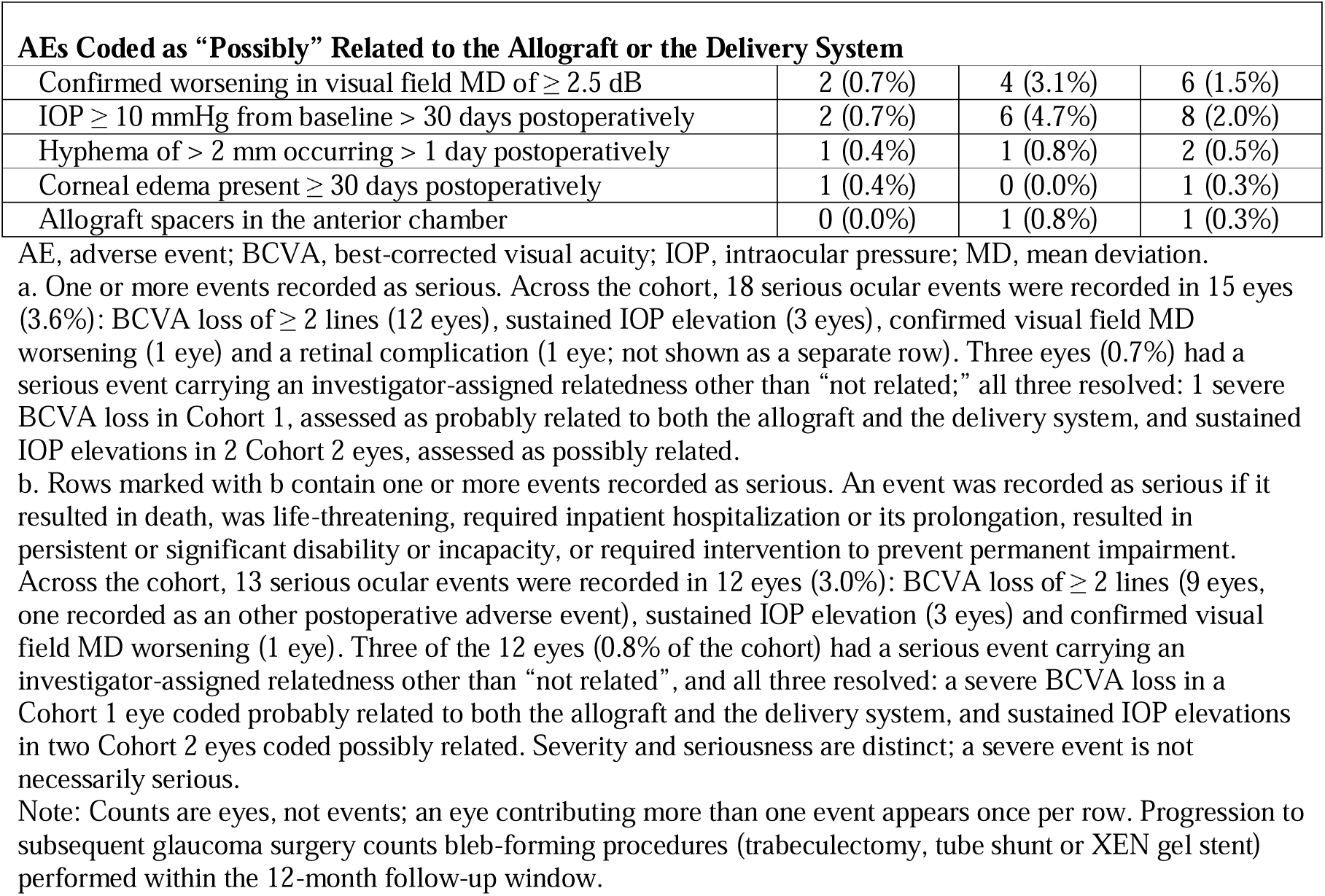
Summary of Ocular Adverse Events (N=395)

| **Ocular AEs of Interest, n (%)** | **Cohort 1<br>(N=267)** | **Cohort 2<br>(N=128)** | **Total<br>(N=395)** |
|---|---|---|---|
| Progression to subsequent glaucoma surgery | 2 (0.7%) | 25 (19.5%) | 27 (6.8%) |
| IOP ≥ 10 mmHg from baseline ≥ 30 days postoperatively<sup>a</sup> | 7 (2.6%) | 27 (21.1%) | 34 (8.6%) |
| BCVA loss of ≥ 2 lines at ≥ 90 days postoperatively<sup>a</sup> | 9 (3.4%) | 6 (4.7%) | 15 (3.8%) |
| Confirmed worsening in visual field MD of ≥ 2.5 dB<sup>a</sup> | 6 (2.2%) | 7 (5.5%) | 13 (3.3%) |
| Macular edema | 7 (2.6%) | 0 (0.0%) | 7 (1.8%) |
| Hyphema of > 2 mm occurring > 1 day postoperatively | 4 (1.5%) | 2 (1.6%) | 6 (1.5%) |
| Clinically significant hypotony<sup>b</sup> | 0 (0.0%) | 1 (0.8%) | 1 (0.3%) |
| Corneal edema present ≥ 30 days postoperatively | 2 (0.7%) | 0 (0.0%) | 2 (0.5%) |
| **AEs Coded as “Definitely” or “Probably” Related to the Allograft or the Delivery System** |  |  |  |
| IOP ≥ 10 mmHg from baseline ≥ 30 days postoperatively | 4 (1.5%) | 2 (1.6%) | 6 (1.5%) |
| Hyphema of > 2 mm occurring > 1 day postoperatively | 3 (1.1%) | 1 (0.8%) | 4 (1.0%) |
| Unplanned ocular surgical reintervention | 2 (0.7%) | 0 (0.0%) | 2 (0.5%) |
| Anterior rotation of ciliary body / IOL displacement | 1 (0.4%) | 0 (0.0%) | 1 (0.3%) |
| BCVA loss of ≥ 2 lines at ≥ 90 days postoperatively (unspecified mechanism) | 1 (0.4%) | 0 (0.0%) | 1 (0.3%) |
| Choroidal folds | 1 (0.4%) | 0 (0.0%) | 1 (0.3%) |
| Slit lamp worsening of ≥ 2 points | 1 (0.4%) | 0 (0.0%) | 1 (0.3%) |
| Corneal endothelial touch | 0 (0.0%) | 1 (0.8%) | 1 (0.3%) |
| **AEs Coded as “Possibly” Related to the Allograft or the Delivery System** |  |  |  |
| Confirmed worsening in visual field MD of ≥ 2.5 dB | 2 (0.7%) | 4 (3.1%) | 6 (1.5%) |
| IOP ≥ 10 mmHg from baseline > 30 days postoperatively | 2 (0.7%) | 6 (4.7%) | 8 (2.0%) |
| Hyphema of > 2 mm occurring > 1 day postoperatively | 1 (0.4%) | 1 (0.8%) | 2 (0.5%) |
| Corneal edema present ≥ 30 days postoperatively | 1 (0.4%) | 0 (0.0%) | 1 (0.3%) |
| Allograft spacers in the anterior chamber | 0 (0.0%) | 1 (0.8%) | 1 (0.3%) |

AE, adverse event; BCVA, best-corrected visual acuity; IOP, intraocular pressure; MD, mean deviation.
a. One or more events recorded as serious. Across the cohort, 18 serious ocular events were recorded in 15 eyes (3.6%): BCVA loss of ≥ 2 lines (12 eyes), sustained IOP elevation (3 eyes), confirmed visual field MD worsening (1 eye) and a retinal complication (1 eye; not shown as a separate row). Three eyes (0.7%) had a serious event carrying an investigator-assigned relatedness other than “not related;” all three resolved: 1 severe BCVA loss in Cohort 1, assessed as probably related to both the allograft and the delivery system, and sustained IOP elevations in 2 Cohort 2 eyes, assessed as possibly related.
b. Rows marked with b contain one or more events recorded as serious. An event was recorded as serious if it resulted in death, was life-threatening, required inpatient hospitalization or its prolongation, resulted in persistent or significant disability or incapacity, or required intervention to prevent permanent impairment. Across the cohort, 13 serious ocular events were recorded in 12 eyes (3.0%): BCVA loss of ≥ 2 lines (9 eyes, one recorded as an other postoperative adverse event), sustained IOP elevation (3 eyes) and confirmed visual field MD worsening (1 eye). Three of the 12 eyes (0.8% of the cohort) had a serious event carrying an investigator-assigned relatedness other than “not related”, and all three resolved: a severe BCVA loss in a Cohort 1 eye coded probably related to both the allograft and the delivery system, and sustained IOP elevations in two Cohort 2 eyes coded possibly related. Severity and seriousness are distinct; a severe event is not necessarily serious.
Note: Counts are eyes, not events; an eye contributing more than one event appears once per row. Progression to subsequent glaucoma surgery counts bleb-forming procedures (trabeculectomy, tube shunt or XEN gel stent) performed within the 12-month follow-up window.

Across both cohorts, 37 ocular adverse events in 31 eyes (7.8%) were assessed by the investigator as possibly, probably, or definitely related to either the allograft or the delivery system. Thirteen events in 12 eyes (3.0%) met the protocol definition of serious.

In one Cohort 2 eye, two of the scleral allograft spacers were found in the anterior chamber during the early postoperative period, having moved forward out of the cyclodialysis cleft. The eye was returned to the operating room, where the spacers were retrieved and repositioned within the cleft. The investigator graded the event moderate in severity and coded it as possibly related to the allograft. The eye recovered without sequelae and required no further intervention. This was the only eye in which allograft tissue moved into the anterior chamber; no other case of allograft migration, dislocation, or extrusion was recorded. No case of corneal endothelial decompensation, endophthalmitis, or sympathetic ophthalmia was recorded across the 395-eye cohort.

IOP elevations (≥ 10 mmHg above baseline, occurring > 30 days postoperatively) were recorded in 34 eyes (8.6%) overall and were distributed unevenly between cohorts: 27 eyes (21.1%) in Cohort 2 compared with 7 (2.6%) in Cohort 1. This disparity reflects the more refractory disease profile of the standalone cohort, which had a higher burden of prior glaucoma procedures and a greater preoperative medication burden. The rate is higher than the 5.0% observed when the same cohort was examined at 6 months, as expected when the observation window is doubled.

Confirmed worsening in visual field mean deviation of ≥ 2.5 dB was recorded in 13 eyes (3.3%), 6 in Cohort 1 (2.2%) and 7 in Cohort 2 (5.5%). BCVA loss of ≥ 2 lines measured at ≥ 90 days postoperatively was recorded in 15 eyes (3.8%), 9 in Cohort 1 (3.4%) and 6 in Cohort 2 (4.7%). Fourteen of these were recorded under the protocol BCVA definition, and one, in a Cohort 1 eye, was recorded as an other postoperative adverse event with the narrative term BCDVA loss; it is counted here because it is a loss of the same magnitude. Investigators coded 14 of the 15 events as unrelated to both the allograft and the delivery system, and one as probably related to both (Table 2). One further event, in a Cohort 2 eye, was recorded during the study and subsequently withdrawn after site query confirmed that the acuity had not been measured with best correction; it is excluded from all counts reported here.

The etiologies recorded in the case narratives for the unrelated BCVA-loss events were: pre-existing ocular comorbidities, disease progression (including epiretinal membrane), non-arteritic ischemic optic neuropathy, and advanced glaucomatous progression, consistent with the advanced-disease characteristics of this cohort. These are investigator-assigned attributions recorded at the time of the event rather than independent adjudications, and in a single-arm study the contribution of the procedure to vision loss in eyes with advanced disease and multiple comorbidities cannot be isolated. In Cohort 1, five of the BCVA loss events ultimately resolved per investigator assessment, while the remainder either persisted or had unresolved or unknown status at the time of analysis. One of the resolved events was attributable to posterior capsular opacification and recovered completely following YAG capsulotomy; the other resolved events recovered with conservative management.

Macular edema was observed in 7 eyes (1.8%), all in Cohort 1 (2.6%), consistent with the known risk of cystoid macular edema following phacoemulsification. Corneal edema persisting beyond 30 days was recorded in two Cohort 1 eyes (0.5%), one of which was coded possibly related to the delivery system; both resolved. One Cohort 2 eye had an event recorded as corneal endothelial touch, graded mild and coded probably related to the allograft, which also resolved. No eye met the protocol definition of corneal endothelial decompensation.

Hyphema > 2 mm that remained after day 1 postoperatively occurred in 6 eyes (1.5%), similarly distributed across cohorts (Cohort 1: 4 eyes, 1.5%; Cohort 2: 2 eyes, 1.6%); all resolved with conservative management, and 6 were investigator-assessed as related, as expected for an intraoperative bleeding event. One case of clinically significant hypotony was observed in a Cohort 2 eye (0.8%; 0.3% of the cohort) that had presented with severe, refractory open-angle glaucoma at baseline (preoperative IOP 32 mmHg on more than five IOP-lowering medications). Postoperative IOP decreased to 6 mmHg by Month 1 and persisted at 3–4 mmHg through Month 12 despite treatment with topical corticosteroids, difluprednate and atropine; the eye remained under active observation at the time of data extraction. The investigator classified this event as not related to the bio-interventional procedure.

Anterior chamber inflammation was uncommon beyond the first postoperative month. At visits ≥ 30 days after surgery, to which every eye contributed at least one graded assessment, cells were recorded in 32 of 395 eyes (8.1%) and reached grade ≥ 2+ in 6 (1.5%). Flare was recorded in 4 eyes (1.0%). Rates were similar in the two cohorts (cells in 7.5% of Cohort 1 and 9.4% of Cohort 2). Prevalence declined over follow-up, from 9.4% of eyes at Month 1 (21 of 224) to 4.1% at Month 6 (14 of 340) and 2.3% at Month 12 (9 of 394), and 18 of the 21 eyes (86%) with cells at Month 1 had none at Month 12. One eye (0.3%) had an adverse event recorded as chronic anterior uveitis, graded mild and coded as unrelated to the allograft and the delivery system.

One case of shallow anterior chamber with iridocorneal apposition (without lens or corneal touch) was reported. This event followed a subsequent tube shunt procedure performed for uncontrolled IOP, was investigator-assessed as not related to the study procedure, and resolved with same-day anterior-chamber washout.

Twenty-seven eyes (6.8%) progressed to secondary incisional glaucoma surgery (trabeculectomy, tube shunt, or XEN gel stent) within the 12-month follow-up window. The overwhelming majority occurred in Cohort 2 (25 eyes, 19.5%), with only two Cohort 1 eyes (0.7%) requiring secondary incisional surgery. Sixteen of the 27 (4.1% of the cohort) had occurred by 6 months, so roughly 40% of the secondary surgeries in this cohort took place in the second half of the first year. This imbalance between cohorts is consistent with the substantially greater preoperative disease burden and higher rate of prior failed glaucoma procedures in the standalone cohort, for whom bio-reinforced cyclodialysis represented a later-line intervention.

### Serious Ocular Adverse Events

Among the 395 eyes included for analysis, 13 serious ocular adverse events were recorded in 12 eyes (3.0%): 4 eyes in Cohort 1 (1.5%) and 8 in Cohort 2 (6.2%). Most serious AEs (10/13) were assessed as unrelated to the allograft or delivery system. Three were assessed as related: one (0.3%) severe loss of BCVA and two (0.5%) sustained IOP elevations.

### Endothelial Cell Safety

Specular microscopy is optional in the CREST Study because it is a real-world protocol and such measurements are not considered part of routine care for glaucomatous eyes. However, endothelial cell counts are recorded when specular microscopy is performed for any reason. Results are summarized in Table 3. A baseline measurement was recorded in 149 of 395 eyes (38%), and at the time of analysis 91 eyes (23%) had paired baseline and month-12 measurements. Among these eyes, mean endothelial cell density decreased from 2,371 to 2,210 cells/mm², a mean change of −6.5% (−8.7%, −4.4%; p < 0.001). The decrease was larger in Cohort 1: density changed −7.6% (−10.0%, −5.2%; p < 0.001; N = 74), whereas in Cohort 2 the change was −1.9% (−6.5% to +2.6%; p = 0.386; N = 17). The between-cohort difference was 5.7 percentage points (95% CI 0.6, 10.7; p = 0.029), though the cohorts differ in lens status and baseline density (2,446 vs 2,044 cells/mm²) and were not randomly selected for specular testing.

**Table 3.** Corneal Endothelial Cell Density from Baseline to Month 12.

| **ECD, cells/mm<sup>2</sup>** | **Cohort 1<br>(Paired N=74)** | **Cohort 2,<br>(Paired N=17)** | **Pooled<br>(Paired N=91)** |
|---|---|---|---|
| Baseline, mean (SD) | 2,446 (389) | 2,044 (611) | 2,371 (462) |
| Month 12, mean (SD) | 2,256 (423) | 2,007 (623) | 2,210 (473) |
| Mean change, % (95% CI) | −7.6% (−10.0%, −5.2%) | −1.9% (−6.5%, +2.6%) | −6.5% (−8.7%, −4.4%) |
| p-value for change | p < 0.001 | p = 0.386 | p < 0.001 |

CI, confidence interval; ECD, endothelial cell density; SD, standard deviation.
Notes: Specular microscopy is optional under the CREST protocol and was performed at the investigators’ discretion. Because testing was elective, these eyes are not a random sample of the cohort, and the direction of selection bias cannot be determined. P-values based on two-sided paired t-tests of the within-eye percent change from baseline, without adjustment for multiple comparisons.

### IOP and Medication Outcomes

At the 12-month assessment, mean medicated IOP decreased from 19.1 mmHg (18.5, 19.7) to 14.5 mmHg (14.1, 14.9), a mean absolute reduction of 4.6 mmHg (3.9, 5.3) and relative reduction of 18.4% (15.6%, 21.2%; p < 0.001). Mean IOP at Month 1 was 15.3 mmHg. At Month 12, 85% of eyes (337/395) achieved IOP ≤ 18 mmHg, and 66% (260/395) achieved IOP ≤ 15 mmHg. Fifty-one percent of eyes (200/395) achieved ≥ 20% IOP reduction from baseline. By cohort, IOP decreased from 18.1 to 14.0 mmHg in Cohort 1 (−4.1% [–4.7%, −3.5%) and from 21.3 to 15.6 mmHg in Cohort 2 (−5.6% [−7.3%, −4.0%).

On the more stringent composite outcome of IOP between 6–18 mmHg without medication increase from baseline, 77.2% of eyes with paired medication and IOP data (305/395) met this target (Cohort 1, 83.5%; Cohort 2, 64.1%).

Among 395 eyes with paired baseline and 12-month medication data, mean medication burden decreased from 1.9 ± 1.5 to 1.2 ± 1.4, a within-eye reduction of 0.71 ± 1.36 medications (p < 0.001). The proportion of eyes on no IOP-lowering medication increased from 18% at baseline to 42% at 12 months (p < 0.001).

## Discussion

This multicenter real-world study represents the largest dataset to date evaluating the safety of bio-interventional uveoscleral outflow enhancement. Through 12 months, no eye developed corneal endothelial decompensation, endophthalmitis, or sympathetic ophthalmia. Allograft tissue moved into the anterior chamber in just one eye and was successfully repositioned without sequelae. Most events were self-limiting or managed conservatively.

The cohort includes both combined cataract and standalone procedures across multiple centers, providing a breadth of evidence that single-center series and small pivotal trials cannot offer.

Three features of the design bound what can be concluded from these data. First, CREST is a single-arm study with no concurrent comparator and no randomization, so contextualization with published literature must be interpreted with caution. Second, seriousness and relatedness were assigned by unmasked treating investigators without independent adjudication, and attribution is therefore subject to bias of unknown direction. Third, the allograft manufacturer funded the database.

### Corneal Endothelial Safety

Endothelial safety is a central question for any supraciliary procedure, because it is the question on which the CyPass supraciliary micro-stent was withdrawn in 2018 [5, 6]. In the present procedure, deployment of the soft, bio-conforming scaffold within the cyclodialysis cleft, without anterior-chamber protrusion, is a design feature intended to limit corneal endothelial exposure.

In understanding the safety implications of ECD changes after bio-reinforced cyclodialysis, both cohorts are clinically relevant and generally go into the procedure with very different baseline disease characteristics and prior interventions. The combined setting (Cohort 1) is more reflective of the state in which most patients would encounter the procedure, while the standalone setting (Cohort 2) is the one in which the allograft acts without the confounding endothelial trauma of phacoemulsification. As expected, the observed loss was larger in the combined cohort, and the between-cohort difference was 5.7 percentage points (95% CI 0.6, 10.7; p = 0.029). The observed 7.6% decline at one year in Cohort 1 lands within the range reported for routine cataract surgery and for the phacoemulsification-only control arms of the MIGS pivotal trials, in which significant endothelial cell loss at five years occurred in 6% to 11% of control eyes [11]. The magnitude seen in Cohort 1 is therefore what cataract surgery alone would be expected to produce, and it is difficult to attribute an additional effect to the allograft on these data.

The change in Cohort 2 did not reach significance, but the confidence interval runs from −6.5% to +2.6% and is therefore compatible with a loss as large as that seen in Cohort 1. The two cohorts also differ in more than the presence of cataract surgery, including lens status, prior surgeries, and baseline endothelial density. Because this is a real-world study, eyes were not randomized or stratified to account for these imbalances. The between-cohort difference is therefore descriptive rather than a controlled estimate of any contribution from the allograft. Additionally, the eyes measured are not a random sample of either cohort, and the direction of the resulting selection bias cannot be determined: sites may have imaged eyes of particular concern, or conversely only those with the equipment and the most routine postoperative courses.

The more important limit is temporal. In the pooled pivotal-trial analysis, endothelial cell loss with the CyPass micro-stent accelerated relative to controls only after the second postoperative year, whereas the Hydrus microstent showed its loss early and then tracked controls [11]. Twelve-month data cannot distinguish between those two trajectories. What the present analysis supports is narrower than an endothelial safety claim: no early signal is apparent in either cohort, and the loss that did occur is of the magnitude and distribution that concurrent cataract surgery would predict. Whether that holds beyond two years is the question the 24-month CREST analysis is designed to answer, and protocol-mandated specular microscopy in a pre-specified subset would strengthen it considerably.

The clinical findings are consistent with the quantitative data: no eye met the protocol definition of corneal endothelial decompensation, persistent corneal edema was recorded in two eyes, and endothelial touch in one. Other supraciliary approaches that use a flexible or conforming form factor have reported more reassuring endothelial findings than the rigid microstent withdrawn in 2018 [12, 13]. However, those are separate studies in separate populations, and we draw no comparison with them here.

### IOP Elevations

The 8.6% rate of protocol-specified IOP elevations (> 30 days postoperatively) is clinically relevant to characterize, and is higher than the 5.0% seen in the same cohort at 6 months simply because the observation window is longer. For context, the 2025 AAO special commentary on MIGS clinical endpoints noted that IOP spikes are among the most commonly reported adverse events across canal-based MIGS, with reported rates ranging from 1% to 33.3% in randomized clinical trials depending on the procedure and the definition used [14]. The present rate falls within that range, which spans more than an order of magnitude and reflects definitional heterogeneity as much as procedural difference; it should not be read as evidence of an advantage. The concentration of these events in the standalone cohort (21.1% versus 2.6%) is the more informative observation, and is consistent with the markedly greater disease burden of eyes referred for a later-line procedure. Most resolved without additional incisional surgery.

On case-level review, 49% of IOP elevation events occurred within the first 90 days postoperatively and 5% within the first 30 days, at a median of 106 days. Roughly half therefore arose in the second half of the first year, a distribution that a 6-month observation window would have captured only partially. Documented mechanisms by site investigators included corticosteroid response (managed by tapering topical corticosteroids), late cleft scarring or membrane formation (managed by laser suture lysis or revision), and IOP elevation in eyes with concurrent risk factors such as intravitreal injection or prior failed surgery. Most events were managed medically; a subset ultimately required incisional secondary surgery, predominantly in eyes with advanced baseline disease.

### Hyphema Rates

The 1.5% rate of hyphema (> 2 mm, occurring > 1 day postoperatively) can be placed alongside published figures from other MIGS categories, with the caveat that these come from different populations and use different hyphema definitions and ascertainment windows. A 2024 systematic review and meta-analysis of 95 studies and 9,733 eyes undergoing combined phacoemulsification and MIGS reported procedure-specific hyphema rates of 27.7% for gonioscopy-assisted transluminal trabeculotomy (GATT), 15.5% for non-GATT goniotomy approaches, and 3.5% for trabecular meshwork bypass procedures [15]. The rate observed here sits at the low end of that published range, which is notable given that the procedure involves instrumentation of the ciliary body, a more vascularized territory than the trabecular meshwork, and may reflect the controlled ab-interno technique that avoids scleral incisions and limits direct ciliary body trauma. In the absence of a concurrent comparator this remains a descriptive observation. All 6 hyphema events resolved with conservative management; the one eye on anticoagulant therapy resolved over approximately 77 days without medication change or surgical intervention.

### Inflammation Rates

Inflammation is a reasonable concern on anatomical grounds, since cyclodialysis separates the ciliary body from the scleral spur and places allograft tissue against uveal tissue. The observed rates were low: beyond the first postoperative month, cells were recorded in 8.1% of eyes and reached grade ≥ 2+ in 1.5%, flare in 1.0%, and prevalence fell to 2.3% by Month 12 with most affected eyes quiet at one year. Unlike the endothelial data, these gradings were captured almost completely, so the estimate does not depend on a selected subset. Two caveats remain: all eyes received a postoperative corticosteroid regimen at the surgeon’s discretion, so these are rates under treatment rather than natural history, and slit lamp grading is a subjective assessment made by unmasked investigators without a concurrent comparator.

### BCVA Loss and Disease Severity

The 3.8% rate of BCVA loss ≥ 2 lines measured at ≥ 90 days requires careful contextual interpretation. Canal-based MIGS trials have generally reported BCVA loss rates of 1–2% in lower-risk elective populations, but these studies typically exclude eyes with significant pre-existing ocular comorbidities and are frequently performed combined with phacoemulsification, which itself improves vision. The CREST population differs in important ways: 24% of eyes were designated as candidates for bleb-forming surgery, reflecting severe or refractory disease; the cohort included pseudophakic eyes in which phacoemulsification-related vision improvement cannot offset procedure-related loss; and enrollment imposes no ceiling on disease severity or comorbidity. Investigators coded 14 of the 15 events as unrelated to the allograft and the delivery system, recording pre-existing epiretinal membrane, non-arteritic ischemic optic neuropathy, advanced glaucomatous field loss and systemic conditions as the etiologies. We report these as the attributions recorded in the study rather than as an independent determination of cause. Because the assignments were made by unmasked treating investigators and no comparator group exists, these data cannot exclude that some portion of the vision loss in this cohort is procedure-related, and the one event coded as related is reported above.

A comprehensive review of MIGS device safety profiled trabecular bypass implants and Schlemm’s canal procedures as generally the safest class of MIGS, with predominantly self-limited, non-vision-threatening complications, while subconjunctival gel stent insertion carries bleb-related and stent-specific adverse events broadly analogous to trabeculectomy [16]. Where bio-reinforced cyclodialysis will ultimately sit in that hierarchy cannot be determined from a single-arm cohort followed for twelve months, and we make no such claim. What these data establish is narrower: across 395 eyes followed for a year, the procedure was not associated with corneal endothelial decompensation, endophthalmitis or sympathetic ophthalmia; allograft tissue entered the anterior chamber in one eye and was repositioned; endothelial cell loss was larger in eyes undergoing concurrent cataract surgery and of the magnitude that cataract surgery alone would predict; and transient IOP elevation, hyphema and secondary surgery occurred at the rates described above. Whether the supraciliary route can be accessed without the late endothelial attrition that emerged with a rigid supraciliary microstent beyond two years remains the central open question for this procedure class, and twelve-month data narrow it without closing it.

### Limitations

Several limitations bound the interpretation of these findings. The first is follow-up duration. Twelve months is longer than the 6-month window in which this cohort was first examined, and long enough to capture the second half of the first postoperative year, during which roughly half of the IOP elevations and 40% of the secondary surgeries in this cohort occurred. It is still shorter than the interval over which the endothelial signal with an earlier supraciliary micro-stent became apparent. Complications with a characteristically delayed onset, including progressive fibrosis, late cleft closure, chronic low-grade inflammation, delayed migration and progressive endothelial cell loss, may still be incompletely captured. CREST follows all enrolled eyes to 24 months, and that analysis remains the relevant test of durability.

Second, the study is single-arm. There is no concurrent comparator and no randomization, so event rates cannot be attributed to the procedure rather than to the underlying disease, to concurrent phacoemulsification, or to the natural history of an advanced-glaucoma population. Published rates from other procedures are cited in the Discussion for orientation only; differences in population, adverse event definition, ascertainment and follow-up make formal comparison inappropriate, and none was performed.

Third, causality assessment was neither masked nor independent. Severity, seriousness and relatedness were assigned by treating site investigators, some of whom have financial relationships with the sponsor, and the sponsor funded the study and maintains the database. No blinded adjudication committee reviewed the events. Attribution of adverse events to pre-existing comorbidity rather than to the procedure is therefore an investigator judgement subject to bias whose direction cannot be established from these data. Readers should weight the event counts, which are objective, more heavily than the relatedness coding, which is not.

Fourth, endothelial cell data remain incomplete and non-randomly obtained. Specular microscopy is optional under the protocol and was performed at investigator discretion, yielding a baseline measurement in 38% of eyes and paired measurements in 23%. The standalone subgroup, on which the interpretation above rests, comprises only 17 eyes. No unbiased estimate of endothelial change can be derived from a sample selected in this way, and the analysis should be read as hypothesis-consistent rather than confirmatory. Protocol-mandated specular microscopy in a pre-specified subset is planned for the longer-term analyses.

Finally, eyes were analyzed at the eye level without adjustment for inter-eye correlation in the 97 patients who contributed both eyes, which may produce modestly narrow confidence intervals; and the January 2026 data export means that eyes enrolled closer to the export date contribute shorter follow-up than those enrolled earlier, so late adverse event capture is less complete in the more recently enrolled eyes.

### Conclusions

In this prospective, multicenter study of 395 eyes followed for at least 12 months, bio-interventional uveoscleral outflow enhancement was not associated with corneal endothelial decompensation, endophthalmitis or sympathetic ophthalmia. Allograft tissue entered the anterior chamber in one eye and was repositioned without sequelae; no other migration, dislocation or extrusion was recorded. Sustained IOP elevation occurred in 8.6% of eyes, hyphema > 2 mm in 1.5% and clinically significant hypotony in 0.3%, and most events were self-limiting or managed medically. In the subset with paired specular microscopy, endothelial cell loss was larger in eyes undergoing concurrent phacoemulsification than in standalone eyes and was of the magnitude that cataract surgery alone would predict, though the standalone subgroup is small and testing was elective. These findings do not establish comparative safety: the study has no control arm and causality was assigned by unmasked treating investigators. Twelve months does not yet reach the interval over which an earlier supraciliary micro-stent diverged from controls, and the 24-month CREST analysis, with protocol-mandated endothelial monitoring, remains the decisive test.

## Author Contributions

Douglas Rhee, James Lewis, Ticiana De Francesco, Kiper Nelson, Tsontcho Ianchulev and Steven R. Sarkisian Jr. contributed to the conception and design of the study. Douglas Rhee, James Lewis, Ticiana De Francesco, Kiper Nelson and Steven R. Sarkisian Jr. acquired the clinical data. Tsontcho Ianchulev and Douglas Rhee analyzed and interpreted the data. All authors drafted or critically revised the manuscript for important intellectual content, approved the version to be published, and agree to be accountable for all aspects of the work.

## Disclosure

The sponsor funded the study, designed the case report forms and maintains the study database. Adverse event severity, seriousness and relatedness were recorded by the treating site investigators; no independent or masked adjudication committee was convened. The authors had full access to the data and take responsibility for the integrity of the analysis and the decision to submit for publication.

## Ethics Statement

The CREST Study was conducted in accordance with the Declaration of Helsinki and received approval from the relevant Institutional Review Boards for all participating investigators (Advarra IRB and site-specific boards).

## Consent

All participants provided written informed consent prior to enrollment.

## Data Availability Statement

The data supporting the findings of this study are held in the CREST Study database by the sponsor, Iantrek, Inc. De-identified data are available from the corresponding author on reasonable request, subject to the sponsor’s approval and an appropriate data sharing agreement.

## Acknowledgments

The authors have no acknowledgments to declare.

## Figure Legends

**1Figure 1. Placement of Allograft Cleft Reinforcement** (Top) Bio-interventional cyclodialysis with the allograft spacer fully deployed to maintain a durable uveoscleral outflow conduit. (Lower-left) processed micro-trephined scleral bio-scaffold composed of porous, acellular, hydrophilic, collagenous sterile matrix. (Lower-right) Sleeved interventional deployment with a cyclodialysis cannula.

**2Figure 2. In-situ Images of the Allograft** Clockwise from upper-left: (1) Gonioscopy view of the allograft spacer during deployment. (2) Postoperative 90-day corneal optical coherence tomography (OCT) section of the cyclodialysis with the in-situ allograft reinforcement demonstrating the homologous biologic nature of the tissue in lieu of conventional implantable hardware. (3) Postoperative 90-day longitudinal OCT imaging of the uveoscleral outflow enhancing bio-scaffold designed to maintain a durable aqueous conduit and filtration reservoir and prevent re-stenosis of the cyclodialysis. (4) External transluminating image demonstrating the visible allograft spacers providing the additive reinforcement at 90 days after the procedure.

